# Effect of delays in concordant antibiotic treatment on mortality in patients with hospital-acquired *Acinetobacter* spp. bacteremia: a 13-year retrospective cohort

**DOI:** 10.1101/2020.04.27.20081513

**Authors:** Cherry Lim, Mo Yin, Prapit Teparrukkul, Maliwan Hongsuwan, Nicholas P.J. Day, Direk Limmathurotsakul, Ben S Cooper

**Affiliations:** Mahidol Oxford Tropical Medicine Research Unit, Faculty of Tropical Medicine, Mahidol University, Thailand; Centre for Tropical Medicine and Global Health, Nuffield Department of Medicine, University of Oxford, United Kingdom; Division of infectious disease, University Medicine Cluster, National University Hospital, Singapore, Singapore; Department of Internal Medicine, Sunpasitthiprasong Hospital, Ubon Ratchathani, Thailand

**Keywords:** empirical antibiotic treatment, patient mortality, *Acinetobacter* spp., bacteremia, causal inference

## Abstract

**Background:** Therapeutic options for multidrug-resistant *Acinetobacter* spp. are limited, and resistance to last resort antibiotics in hospitals is increasing globally. Quantifying the impact of delays in concordant antibiotic treatment on patient mortality is important for designing hospital antibiotic policies.

**Methods:** We included patients with *Acinetobacter* spp. hospital-acquired bacteremia (HAB) in a hospital in Thailand over a 13-year period. For each day of stay following the first positive blood culture we considered antibiotic treatment to be concordant if the isolated organism was susceptible to at least one antibiotic given. We used marginal structural models with inverse-probability weightings to determine the association between delays in concordant treatment and 30-day mortality.

**Results:** Between January 2003 and December 2015, 1,203 patients had HAB with *Acinetobacter* spp., of which 682 patients (56.7%) had one or more days of delay in concordant treatment. These delays were associated with an absolute increase in 30-day mortality of 6.6% (95% CI 0.2%-13.0%), from 33.8% to 40.4%. Crude 30-day mortality was substantially lower in patients with three or more days of delays in concordant treatment compared to those with one to two days of delays. Accounting for confounders and immortal time bias resolved this paradox, and showed similar 30-day mortality for one, two and three or more days of delays.

**Conclusions:** Delays in concordant antibiotic treatment were associated with a 6.6% absolute increase in mortality among patients with hospital-acquired *Acinetobacter* spp. bacteremia. If this association is causal, switching fifteen patients from discordant to concordant initial treatment would be expected to prevent one death.

**Funding:** The Mahidol Oxford Tropical Medicine Research Unit (MORU) is funded by the Wellcome Trust [grant number 106698/Z14/Z]. CL is funded by a Wellcome Trust Research Training Fellowship [grant number 206736/Z/17/Z]. MY is supported by a Singapore National Medical Research Council Research Fellowship [grant number NMRC/Fellowship/0051/2017]. BSC is funded by the UK Medical Research Council and Department for International Development [grant number MR/K006924/1]. DL is funded by a Wellcome Trust Intermediate Training Fellowship [grant number 101103]. The funder has no role in the design and conduct of the study, data collection, or in the analysis and interpretation of the data.

## Introduction

Hospital-acquired bloodstream infection related to multidrug-resistant (MDR) *Acinetobacter* spp. is associated with increased patient mortality, especially in developing countries. The proportion of hospital-acquired *Acinetobacter* spp. bacteremias that are MDR can be as high as 75%, and attributable mortality has been estimated to range from 18–41% in developing countries.^1–5^ In a previous study conducted in Thailand, around 15,000 excess deaths per year were estimated to be related to MDR *Acinetobacter* spp..^2^ Therapeutic options for treating MDR *Acinetobacter* spp. infections are limited. Carbapenem, colistin, and tigecycline are currently the last resort antibiotics for drug-resistant *Acinetobacter* spp. bloodstream infection, and increasing resistance to these antibiotics has been reported in developing countries.^6,7^ The spread of resistant pathogens can be fueled by the overuse of broad-spectrum antibiotics in hospital settings,^8^ and *Acinetobacter* spp. has an ability to assemble different mechanisms of resistance.^9^ The World Health Organization recommends that hospital antibiotic policies are developed based on local evidence, with the aims of minimizing morbidity and mortality due to infections, preserving the effectiveness of antimicrobial agents for treatment purposes, and preventing the spread of microbial infections.^10^ A quantitative understanding of the impact of delays in concordant antibiotic treatment on patient mortality among those with hospital-acquired *Acinetobacter* spp. bloodstream infection is important for designing hospital antibiotic policies in areas with a high incidence of drug-resistant *Acinetobacter* spp. infection.

Randomized controlled trials are the “gold standard” design to study the causal relationship between a treatment and an outcome. However, it would be unethical to intentionally delay the provision of concordant antibiotic treatment to a patient. Therefore, an observational cohort study is likely to be the best way to determine the impact of delays in concordant antibiotic treatment on patient outcomes. There are, however, intrinsic limitations with such an approach. Firstly, hospitalized patients vary in severity of underlying illness, which in turn could affect antibiotic prescription behavior and the probability of acquiring drug-resistant infections. At the same time, severity of underlying illness is an important determinant of mortality. This suggests severity of underlying illness is a key confounder to consider when studying the causal relationship between antibiotic treatment and patient in-hospital mortality, and should be adjusted for in the analysis. However, measures of patient severity are not routinely collected in most resource-limited hospital settings. Secondly, antibiotic treatment may change over the course of an infection, and these changes will often be related to the time-varying clinical severity of the patient, which could be a confounder for the current treatment and a mediator for the future treatment (Box 1). Finally, patients may die or be discharged from the hospital before the empirical treatment is completed, and excluding these patients from the final analysis could introduce immortal time bias.^11–13^

#### Box 1. A directed acyclic graph (DAG) to illustrate the definition of confounder and mediator

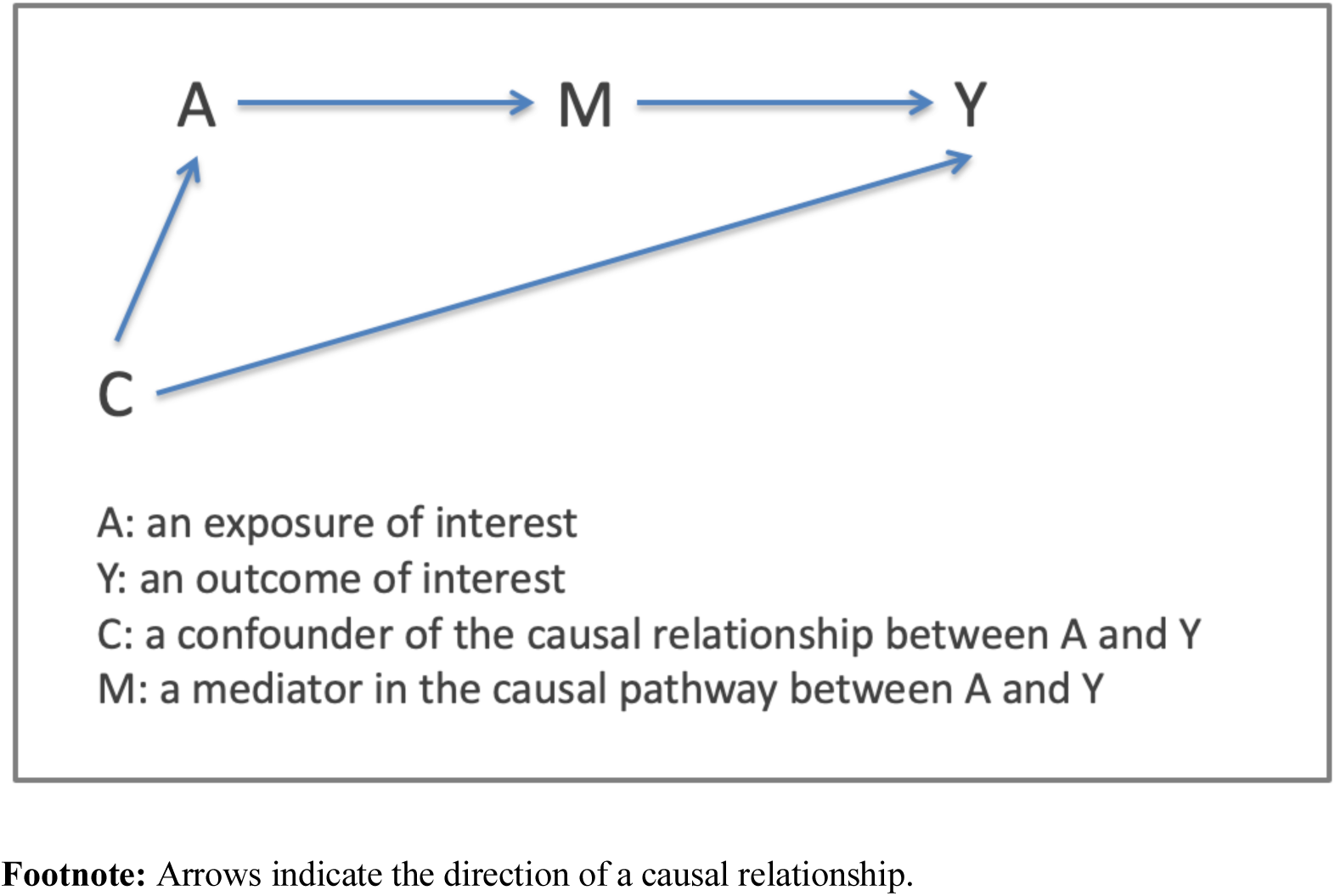

In this study, our objective is to study the relationship between days of delay in concordant antibiotic treatment and patient mortality by performing an analysis on observational data accounting for these distorting factors, aiming, as far as possible, to emulate a randomized trial.

## Methods

### Study design and participants

We identified, from a 13-year retrospective cohort, patients with hospital-acquired bacteremia (HAB) related to *Acinetobacter* spp. in Sunpasitthiprasong Hospital, Thailand. This is a provincial hospital with 1,500 beds. The hospital has a microbiology laboratory that performs culture and isolate identification and antibiotic susceptibility tests on a daily basis. The number of blood cultures performed in 2003 was 11,584 and in 2015 was 56,719. Patients were eligible for inclusion in this study if they had stayed in the hospital longer than 2 calendar days when a blood sample with growth of *Acinetobacter* spp. was collected. During the study period, bacterial culture was performed using standard methodologies for bacterial identification and susceptibility testing based on guidelines of the Bureau of Laboratory Quality and Standards, Ministry of Public Health, Thailand.^14^ Antimicrobial susceptibility was determined using the disk diffusion method based on Clinical and Laboratory Standards Institute (CLSI) guidelines.^15^ The first episode of *Acinetobacter* spp. bloodstream infection (BSI) per eligible patient was included in the analysis. If more than one isolate of *Acinetobacter* spp. with different susceptibility profiles was identified, only the isolate resistant to the largest number of antibiotics tested was included in the analysis. Hospital admission and microbiology databases were merged to identify the cohort of patients. Data on antibiotic prescription, ICD 10 diagnosis, and demographic information of this cohort were then collected for analysis.

The study was approved by the Institutional Review Board of Sunpasitthiprasong Hospital. STROBE recommendations were followed (Supplementary 1).

### Exposure groups

We considered any antibiotics prescribed within three days of the first blood sample collected and with *Acinetobacter* spp. isolated to represent empirical treatment. This is because microbiological identification and antibiotic susceptibility testing usually require three days. An antibiotic regimen was defined as concordant if susceptibility testing indicated that the isolated organism was susceptible to at least one of the antibiotics given. Otherwise the regimen was defined as discordant. Concordance of the antibiotic treatment was determined for each eligible patient on the day of blood sample collection (t=0), one calendar day after blood sample collection (t=1), and two calendar days after blood sample collection (t=2).

### Outcomes

The primary outcome is in-hospital all-cause mortality within 30 days of the first blood sample collected and with *Acinetobacter* spp. isolated. If a patient was discharged alive from the hospital before Day 30 or remained in the hospital on Day 30, then the patient was considered to have survived in the primary analysis.

### Covariate selection

Potential confounders were identified using a directed acyclic graph to represent the presumed causal relationships between antibiotic treatment and patient mortality (Supplementary 2).^16^ The key potential baseline confounders identified were severity of underlying illness,^17^ antibiotic resistance pattern of the *Acinetobacter* spp. isolated from the blood sample, year in which the blood samples were collected, and specialty of the attending physician. We used the time between date of admission and date of blood sample collection, admission to intensive care unit (ICU) on the day of hospital admission, the number of days on antibiotic treatment prior to blood sample collection, and age-stratified Charlson comorbidity index (CCI) score as surrogates of severity of underlying illness. The CCI scores were calculated from the ICD 10 codes given to each patient by the attending physicians.^18^ MDR *Acinetobacter* spp. was defined as previously described.^19^ As data on specialty of the attending physician is not routinely collected in the electronic record, we used the department in which the patient was treated on the day of blood collection as a proxy variable. A time-varying confounder that could affect changes in empirical antibiotic treatment post blood sample collection is severity of infection, which could be affected by the history of treatment and itself may influence the decision on future treatment. The prescription of a vasopressor and transfer to ICU during the infection within the analysis time period were used to represent severity of the infection and both coded as binary time varying variables. Patient demographic information, age and gender, were also included as covariates.

### Statistical analysis

The effects of delays in concordant empirical antibiotic treatment on 30-day mortality were estimated using marginal structural models.^20^ We performed two analyses. The first analysis was to evaluate the impact of one or more days of delays in concordant antibiotic treatment. A propensity score for each patient was calculated to represent the probability of being prescribed with concordant antibiotic on the day of blood collection using a logistic regression model adjusting for the pre-specified confounders and using fractional polynomials to account for non-linear relationships. The propensity scores were then used to calculate stabilized inverse probability weights (IPW), which were applied to a marginal structural model. The second analysis was to evaluate the effect of one-day, two-day, and three or more days of delays in concordant antibiotic treatment. We applied two sets of IPWs to a marginal structural model. Firstly, a propensity score for each patient was calculated to represent the probability of being prescribed with a concordant antibiotic treatment on the day of blood sample collection, one day after, and two days after. The propensity scores were then used to calculate stabilized IPWs. Secondly, to emulate a randomized controlled trial with treatment regimen assigned on enrolment, patients who were discharged or died before completing the treatment were treated as censored observations, and propensity scores were calculated for being uncensored in the cohort.^13^ We then applied a marginal structural logistic regression model with the (multiplicative) total stabilized IPWs to estimate the marginal probability of 30-day mortality under each treatment regimen. Statistical analyses were done using STATA, version 15.1 (StataCorp LP, College station, Texas, USA). All analysis code is provided in Supplementary 3. A simulation study was also performed and confirmed that the procedure could recover the expected 30-day mortality associated with delays in concordant antibiotic treatment.

It is a common practice in Thailand for moribund patients to be discharged and to die at home. This may cause mis-classification of outcomes. To address the issue, we performed a sensitivity analysis (Supplementary 5) and patients who were either discharged without improvement or who rejected treatment and discharged were classified into the group assumed to have died within 30 days.

### Role of the funding source

The funders of the investigators and study had no role in the study design, data collection, data analysis, data interpretation, or writing of the manuscript. CL, DL, and PT had full access to the data described in the study. The corresponding author has the final decision to submit for publication.

## Results

### Patients

Between January 1, 2003, and December 31, 2015, 1,203 patients had a blood sample collected yielding *Acinetobacter* spp after two days of hospitalization (Figure 1). Amongst the eligible patient cohort, 521 patients had no delays in concordant antibiotic treatment (i.e. patients had concordant treatment on the day of blood sample collection); 224 patients had a one-day delay in concordant antibiotic treatment; 119 patients had a two-day delay in concordant antibiotic treatment; and 339 patients had three or more days of delays in concordant antibiotic treatment.

**Figure 1.**
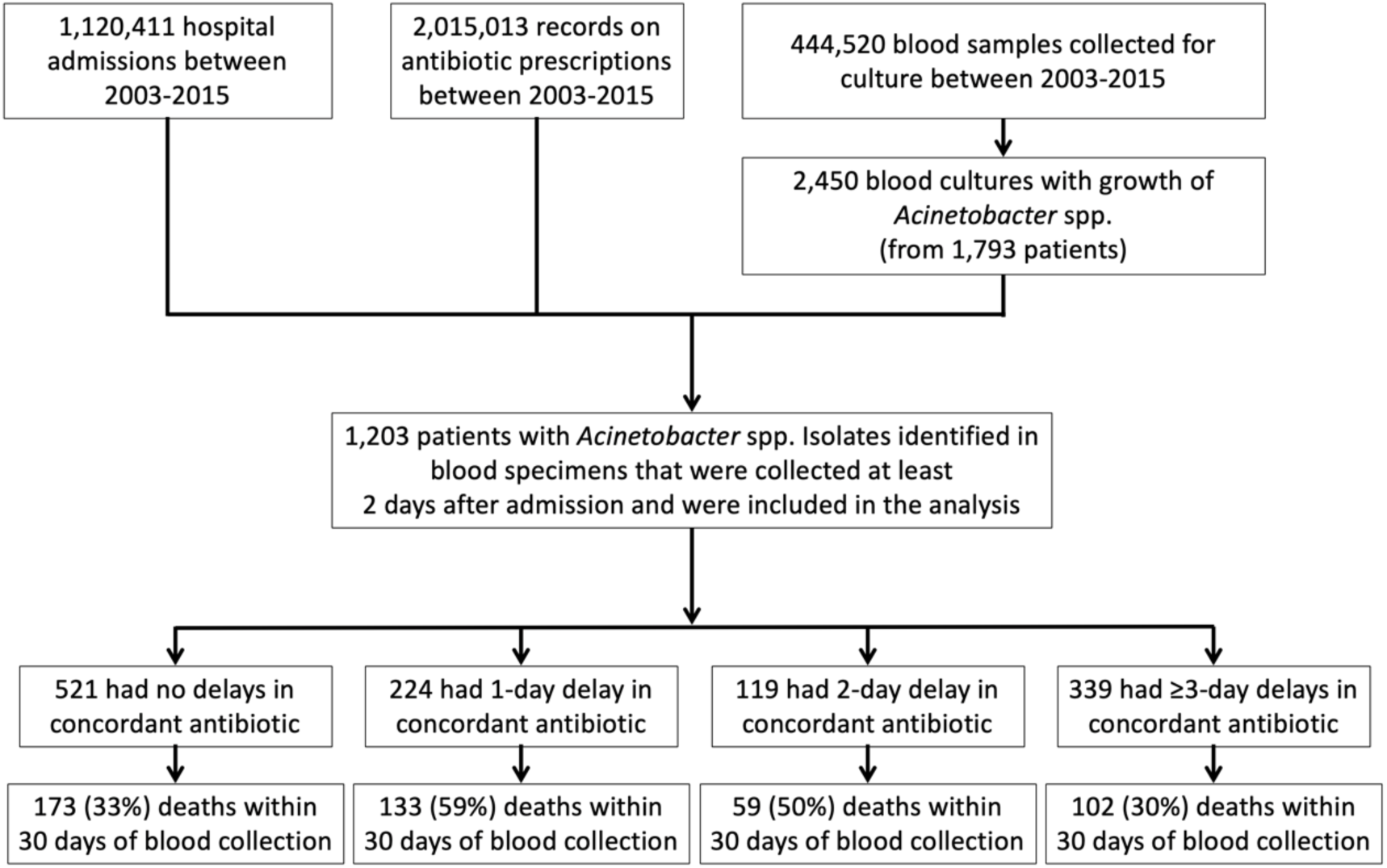
Flow chart of patients identified in the hospital microbiology database and included in the analysis.

Patient characteristics varied across the four groups of exposures (Table 1). The proportion of patients admitted to ICU wards on the day of hospitalization was highest among those having a one-day delay in concordant antibiotic treatment (159 of 224 patients; 71.0%), followed by those having a two-day delay (72 of 119 patients, 60.5%). The proportion of patients with MDR *Acinetobacter* spp. isolates was highest among those who had a one-day delay in concordant antibiotic treatment (206 of 224; 92.0%), followed by those who had three or more days of delay (302 of 339 patients; 89.1%). The cumulative days on antibiotics prior to blood collection also varied across the four groups of patients. The patients who had no delays in concordant antibiotic treatment were on antibiotics the longest prior to blood sample collection, with a median of 8 (IQR: 5–15) days.

**Table 1.**
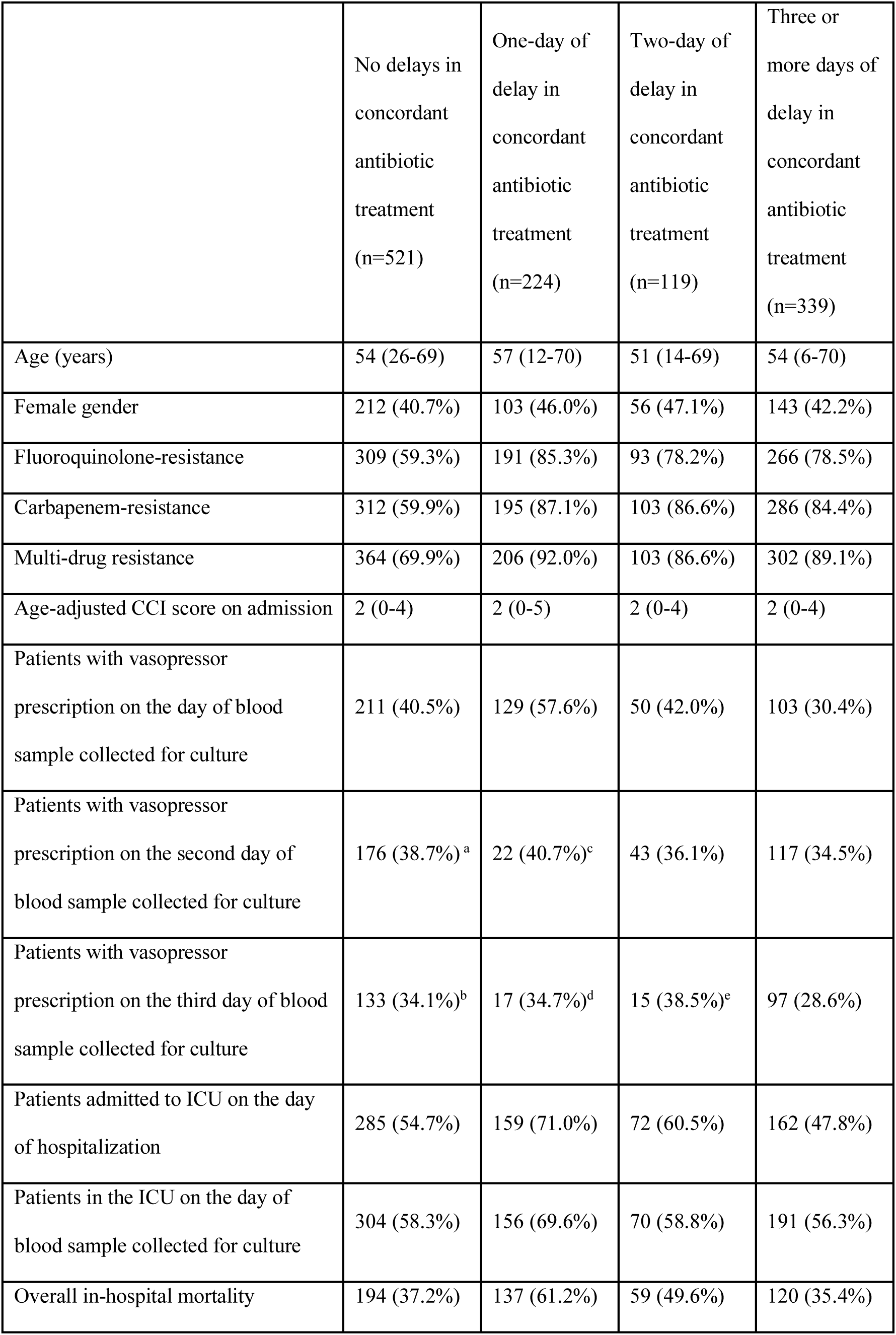
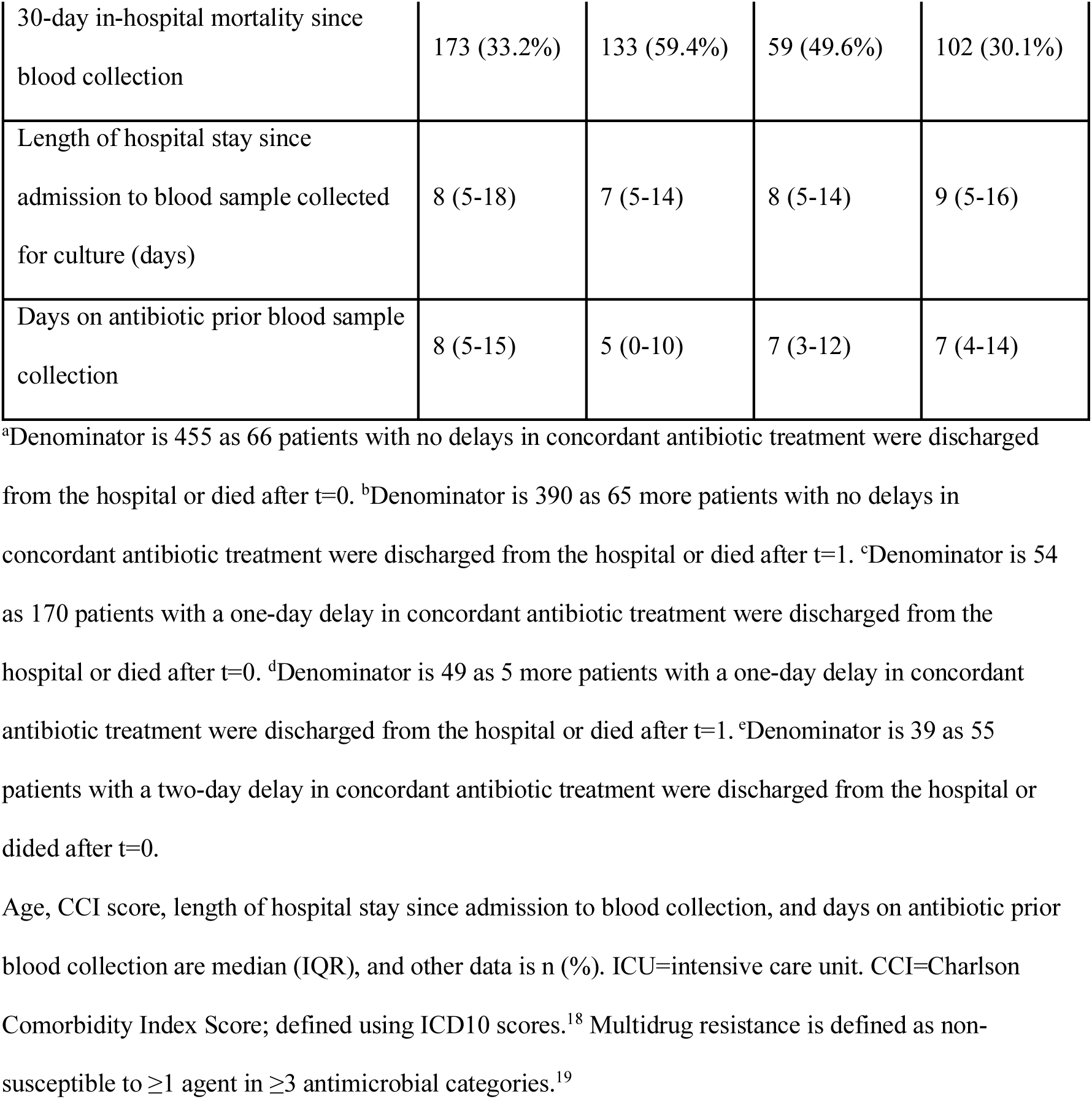
Characteristics of patients with hospital-acquired bloodstream infection related to *Acinetobacter spp.*.

|  | No delays in concordant antibiotic treatment (n=521) | One-day of delay in concordant antibiotic treatment (n=224) | Two-day of delay in concordant antibiotic treatment (n=119) | Three or more days of delay in concordant antibiotic treatment (n=339) |
| --- | --- | --- | --- | --- |
| Age (years) | 54 (26-69) | 57 (12-70) | 51 (14-69) | 54 (6-70) |
| Female gender | 212 (40.7%) | 103 (46.0%) | 56 (47.1%) | 143 (42.2%) |
| Fluoroquinolone-resistance | 309 (59.3%) | 191 (85.3%) | 93 (78.2%) | 266 (78.5%) |
| Carbapenem-resistance | 312 (59.9%) | 195 (87.1%) | 103 (86.6%) | 286 (84.4%) |
| Multi-drug resistance | 364 (69.9%) | 206 (92.0%) | 103 (86.6%) | 302 (89.1%) |
| Age-adjusted CCI score on admission | 2 (0-4) | 2 (0-5) | 2 (0-4) | 2 (0-4) |
| Patients with vasopressor prescription on the day of blood sample collected for culture | 211 (40.5%) | 129 (57.6%) | 50 (42.0%) | 103 (30.4%) |
| Patients with vasopressor prescription on the second day of blood sample collected for culture | 176 (38.7%)<sup>a</sup> | 22 (40.7%)<sup>c</sup> | 43 (36.1%) | 117 (34.5%) |
| Patients with vasopressor prescription on the third day of blood sample collected for culture | 133 (34.1%)<sup>b</sup> | 17 (34.7%)<sup>d</sup> | 15 (38.5%)<sup>e</sup> | 97 (28.6%) |
| Patients admitted to ICU on the day of hospitalization | 285 (54.7%) | 159 (71.0%) | 72 (60.5%) | 162 (47.8%) |
| Patients in the ICU on the day of blood sample collected for culture | 304 (58.3%) | 156 (69.6%) | 70 (58.8%) | 191 (56.3%) |
| Overall in-hospital mortality | 194 (37.2%) | 137 (61.2%) | 59 (49.6%) | 120 (35.4%) |
| 30-day in-hospital mortality since blood collection | 173 (33.2%) | 133 (59.4%) | 59 (49.6%) | 102 (30.1%) |
| Length of hospital stay since admission to blood sample collected for culture (days) | 8 (5-18) | 7 (5-14) | 8 (5-14) | 9 (5-16) |
| Days on antibiotic prior blood sample collection | 8 (5-15) | 5 (0-10) | 7 (3-12) | 7 (4-14) |

<sup>a</sup>Denominator is 455 as 66 patients with no delays in concordant antibiotic treatment were discharged from the hospital or died after t=0. <sup>b</sup>Denominator is 390 as 65 more patients with no delays in concordant antibiotic treatment were discharged from the hospital or died after t=1. <sup>c</sup>Denominator is 54 as 170 patients with a one-day delay in concordant antibiotic treatment were discharged from the hospital or died after t=0. <sup>d</sup>Denominator is 49 as 5 more patients with a one-day delay in concordant antibiotic treatment were discharged from the hospital or died after t=1. <sup>e</sup>Denominator is 39 as 55 patients with a two-day delay in concordant antibiotic treatment were discharged from the hospital or died after t=0.
Age, CCI score, length of hospital stay since admission to blood collection, and days on antibiotic prior blood collection are median (IQR), and other data is n (%). ICU=intensive care unit. CCI=Charlson Comorbidity Index Score; defined using ICD10 scores.<sup>18</sup> Multidrug resistance is defined as non-susceptible to $\geq 1$ agent in $\geq 3$ antimicrobial categories.<sup>19</sup>

The most commonly prescribed antibiotics on the day of blood sample collection were carbapenems (n=312) followed by ceftazidime (n=121). Of 467 patients who died within 30 days after blood sample collection, 294 patients did not have a concordant antibiotic prescription on the day of blood sample collection. Of those patients, 33.7% (99/294) had a prescription of a carbapenem, 16.0% (47/294) did not have an antibiotic prescription, and 9.9% (29/294) had a prescription of a third-generation cephalosporin. Of 736 patients who survived for at least 30 days after the first positive blood sample was collected, 24.2% (178/736) had a prescription of a carbapenem on the day of blood sample collection, 17.1% (126/736) had a prescription of a third-generation cephalosporin, and 10.1% (74/736) did not have an antibiotic prescription.

### Antibiotic treatment concordance on the day of blood sample collection and 30-day in-hospital mortality

Receiving concordant antibiotic treatment on the day of blood collection was associated with reduced 30-day mortality, after adjusting for the pre-specified confounders. Patients given concordant antibiotic treatment on the day of blood collection had an expected 30-day mortality of 33.8% (95% CI 29.1%-38.5%), compared with an expected 30-day mortality of 40.4% (95% CI 36.0%-44.7%) in those not treated with concordant antibiotics. The absolute difference was 6.6% (95% CI 0.2%-13.0%).

### Days of delays in concordant antibiotic treatment and 30-day in-hospital mortality

The crude 30-day in-hospital all-cause mortality was highest among those with a one-day delay in concordant antibiotic treatment (133 of 224 patients; 59.4%), and lowest among those with three or more days of delays in concordant antibiotic treatment (102 of 339 patients; 30.1%) [Table 1]. The discharge pattern of patients under different treatment groups varied over the three days of the initial treatment period (Figure 2). Amongst the 1,203 eligible patients, 236 (19.6%) patients either died or were discharged from the hospital one day after blood was collected for culture and, of those patients, 63.6% died within the hospital.

**Figure 2.**
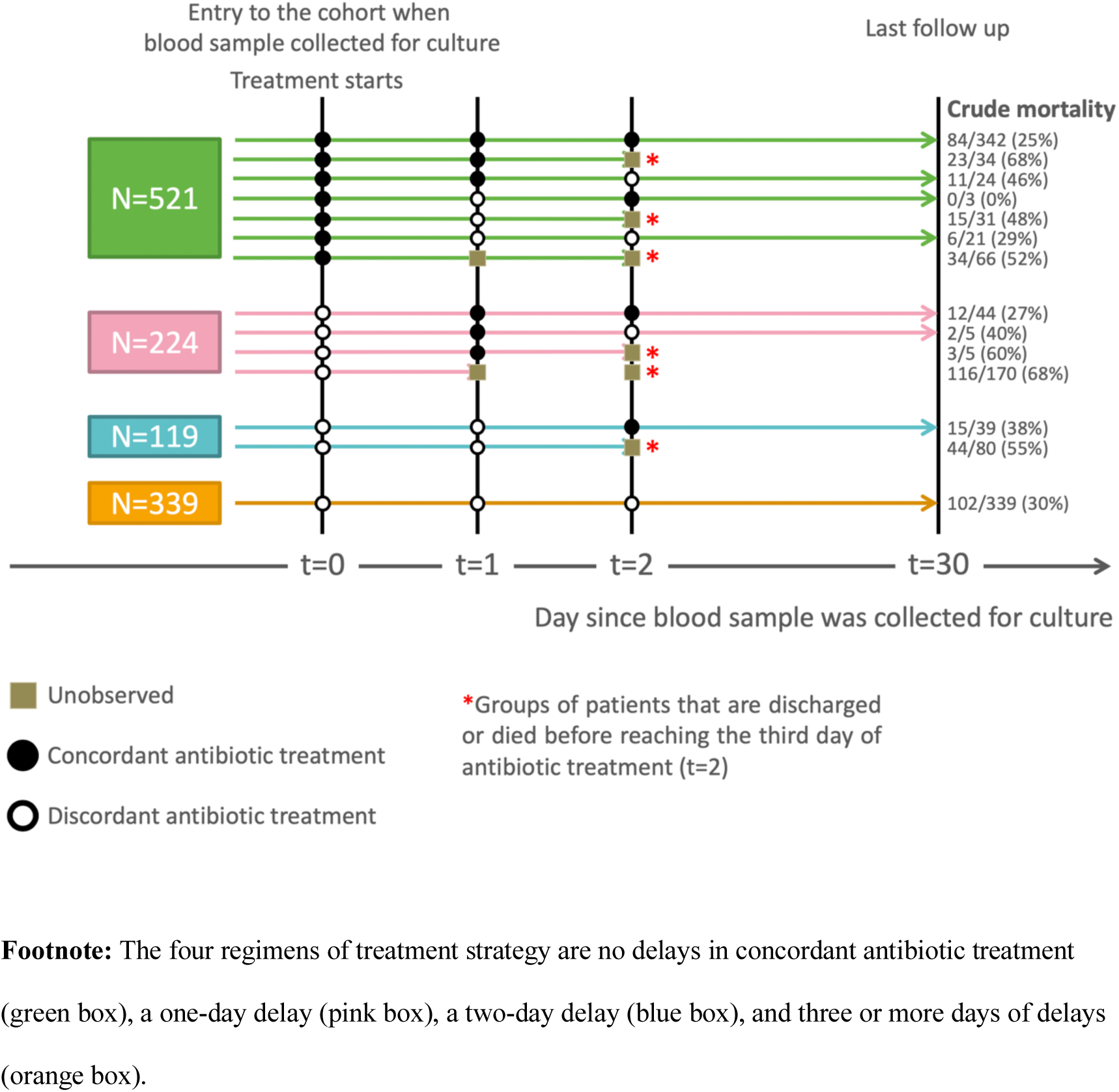
Regimen assignment, all-cause 30-day in-hospital mortality, and discharge patterns over three days post blood sample collection of the study cohort.

The marginal structural model adjusting for baseline confounders, time-varying confounders, and immortal time bias resolved paradoxical observations in the crude data (Table 2). While the crude analysis suggested that patients with three or more days of delays in concordant antibiotic treatment had the lowest mortality, the adjusted analysis found that the expected mortality was the lowest if the patients had no delays, though found no evidence of increasing mortality with increasing delays. However, uncertainty was large. Absolute differences in mortality between no delays in concordant antibiotic treatment and a one-day delay, a two-day delay, and three or more days of delays were 3.0% (95% CI - 12.0%-18.0%); 11.3% (95% CI -3.0%-25.6%); and 1.1% (95% CI -7.8%-10.0%), respectively.

**Table 2.**
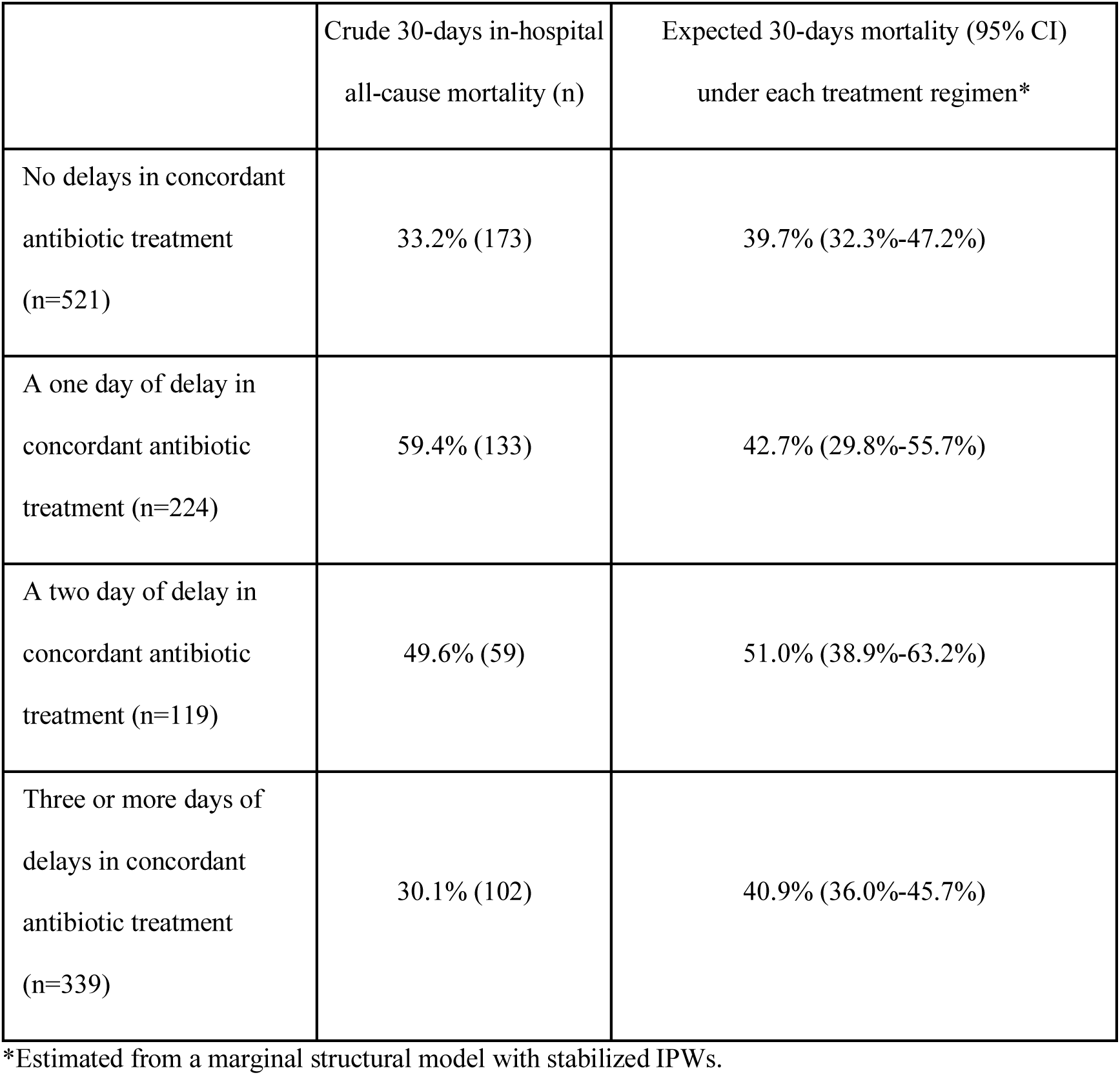
Estimated probability of 30-day mortality under each exposure group.

|  | Crude 30-days in-hospital<br>all-cause mortality (n) | Expected 30-days mortality (95% CI)<br>under each treatment regimen* |
| --- | --- | --- |
| No delays in concordant<br>antibiotic treatment<br>(n=521) | 33.2% (173) | 39.7% (32.3%-47.2%) |
| A one day of delay in<br>concordant antibiotic<br>treatment (n=224) | 59.4% (133) | 42.7% (29.8%-55.7%) |
| A two day of delay in<br>concordant antibiotic<br>treatment (n=119) | 49.6% (59) | 51.0% (38.9%-63.2%) |
| Three or more days of<br>delays in concordant<br>antibiotic treatment<br>(n=339) | 30.1% (102) | 40.9% (36.0%-45.7%) |

### Sensitivity analysis

Under the assumption that patients discharged within 30 days of the first positive blood culture either without improvement or having rejected treatment died within 30 days, a similar effect of delayed concordant treatment was observed. If patients were given a concordant antibiotic treatment on the day of blood sample collection, the expected marginal probability of developing a detrimental outcome (death or discharge without improvement) would be 58.8% (95% CI 53.8%-63.9%), which is lower than if they were not given a concordant antibiotic treatment (62.0% [95% CI 57.7%-66.4%]). The difference was 3.2% (95% CI -3.5%-9.9%). The estimated marginal probabilities of developing a detrimental outcome (death or discharge without improvement) within 30 days of blood collection were 64.6% (95% CI 56.8%-72.4%), 63.2% (95% CI 50.4%-75.9%), 68.5% (95% CI 56.1%-80.8%), 63.4% (95% CI 58.5%-68.3%) for no delays, a one-day delay, a two-day delay, and three or more days of delays in concordant antibiotic treatment respectively (Supplementary 5). The estimated impacts of the treatment regimens on detrimental outcomes were similar to the effects on 30-day in-hospital mortality (Supplementary 6).

## Discussion

After adjusting for measured confounders, we found that delays in concordant antibiotic treatment of one or more days were associated with an absolute increase of 6.6% in 30-day mortality from 33.8% to 40.4%. If this increase is caused by the delay in concordant treatment, it would indicate that we would need to switch empirical antibiotic treatment from discordant to concordant in fifteen patients in order to prevent one 30-day death. There was no evidence of a dose response relationship between number of days of delays in concordant antibiotics and 30-day mortality.

The immortal time bias observed in this cohort is a common phenomenon, and needs to be considered in any study comparing treatment regimens where the observed treatment durations vary.^11,13^ In this study empirical antibiotic treatment over a period of three days after blood collection was considered and only patients who survived up to three days after blood was collected could be classified into the group of “≥3 days of delays in concordant antibiotic treatment”. Hence, by definition, they cannot have died within the first 3 days and for this time period they are effectively “immortal”. This bias will tend to make them appear to survive longer compared to the reference group (no delays in concordant antibiotic treatment). This is reflected in the paradoxical observation that the crude all-cause 30-day mortality was lowest among patients with ≥3 days of delays in concordant antibiotic treatment. Several analytical approaches have been described to adjust for immortal time bias.^11^ For example, an analysis on 964 patients with *Staphylococcus aureus* bacteremia in hospitals in Germany used multivariable Cox regression model with an interaction term of exposure and time to correct for immortal time bias and to study the impact of combined antibiotic therapy on patient survival.^21^ However, even in the absence of unmeasured confounding, causal interpretations of hazard ratios are not stratightforward.^22^

Previous analyses have evaluated the impact of delayed antibiotic treatment on outcomes for patients with bacteremia related to *Acinetobacter* spp., but appropriate adjustment for both time-varying and immortal time biases has been lacking.^23–30^ In a retrospective study on 1,423 patients in hospitals in the United States, pneumonia and/or sepsis patients who received inappropriate empiric therapy (defined as no antibiotic treatment covering the organism administrated within 2 days of the first positive culture) had increased mortality (adjusted relative risk ratio was 1.85 [95% CI 1.35–2.54]).^30^ The study included patients with both community-acquired and healthcare-associated infections; patients with fewer than 2 days of hospitalization were excluded from the analysis. A generalized logistic regression model adjusting for patient demographics, comorbidities, severity of illness, and infection origin was used.^30^ A study in Taiwan on 252 patients with ICU-acquired bloodstream infections caused by *Acinetobacter baumannii* suggested appropriate antibiotic therapy (defined as administration of at least one antibiotic treatment that is appropriate in type, route and dosage within 48 hours of bacteremia onset) reduced 14-day mortality (adjusted odds ratio was 0.22 [95% CI 0.10–0.50]).^27^ The analysis used a multivariable logistic regression model adjusted for APACHE II score measured two days prior to bacteremia onset and malignancy. In this study, among those with APACHE II score >35 more than 70% of patients died within 24 hours in the inappropriate antibiotic group and in the appropriate treatment group no patient died within the initial 48 hours treatment period.^27^ Some of the reported differences in survival probability are therefore expected to be due to immortal-time biases.

In most previous studies antibiotic use has been considered as a binary variable and switching of antibiotic regimens due to changes in clinical symptoms has been neglected.^31^ In hospitals in resource-limited settings, antibiotics are sometimes prescribed even before a clinical specimen is taken for culture, and switching regimen in response to severity of infection is common.^31^ This change in regimen determined by clinical symptoms, if not adjusted for using appropriate methods for time-varying confounders, may also lead to biases.^20^ Marginal structural models have been used to adjust for time-varying confounders in a previous study of the association between appropriate antibiotic treatment for bacteremia on the day the blood culture was taken and mortality and discharge.^32^ The previous study found no evidence for a protective effect of appropriate empirical antibiotic treatment on mortality and discharge, but confidence intervals were wide.^32^ Differences in bacterial species considered, patient characteristics and clinical setting make direct comparison with the current study inappropriate.

Our study has limitations. Firstly, data on severity of infections were not routinely collected, and this is typically the case in LMIC settings. We used admission to ICU and prescription of vasopressor as proxy variables for the severity of infection. These proxy variables will only imperfectly represent the severity of infection and residual confounding is to be expected; however, both are specific in representing severe clinical conditions. Secondly, despite the relatively large sample size, the power to detect a dose-response relationship in the four regimens under evaluation might be low. This is reflected in the wide confidence intervals in our expected mortality under each regimen. Hence, although we do not find evidence of a dose response relationship between delays in concordant treatment and mortality, the results do not rule out the possibility.

In conclusion, we observed a 6.6% absolute increase in mortality among patients with hospital-acquired *Acinetobacter* spp. bacteremia when concordant antibiotic treatment was delayed for one or more days. Accounting for confounding and immortal time biases is necessary when attempting to estimate causal effects of delayed concordant treatment and, in this case, helped resolve paradoxical results in crude data.

## Data Availability

The data that support the findings of this study are available from Sunpasitthiprasong Hospital. Restrictions apply to the availability of these data, which were used under license for this study. Data are available from the authors only with the permission of Sunpasitthiprasong Hospital. STATA code used is provided.

## Notes

### Competing Interest Statement

The authors have declared no competing interest.

